# Impact of baseline SARS-CoV-2 antibody status on syndromic surveillance and the risk of subsequent Covid-19 – a prospective multicentre cohort study

**DOI:** 10.1101/2021.06.09.21258422

**Authors:** Philipp Kohler, Sabine Güsewell, Marco Seneghini, Thomas Egger, Onicio Leal, Angela Brucher, Eva Lemmenmeier, J. Carsten Möller, Philip Rieder, Markus Ruetti, Reto Stocker, Danielle Vuichard-Gysin, Benedikt Wiggli, Ulrike Besold, Stefan P. Kuster, Allison McGeer, Lorenz Risch, Andrée Friedl, Pietro Vernazza, Christian R. Kahlert

## Abstract

**Objectives:** In a prospective healthcare worker (HCW) cohort, we assessed the risk of SARS-CoV-2 infection according to baseline serostatus.

**Methods:** Baseline serologies were performed among HCW from 23 Swiss healthcare institutions between June and September 2020, before the second COVID-19 wave. Participants answered weekly electronic questionnaires covering information about nasopharyngeal swabs (PCR/rapid antigen tests) and symptoms compatible with Coronavirus Disease 2019 (COVID-19). Screening of symptomatic staff by nasopharyngeal swabs was routinely performed in participating facilities. We compared numbers of positive nasopharyngeal tests and occurrence of COVID-19 symptoms between HCW with and without anti-nucleocapsid antibodies.

**Results:** A total of 4’818 HCW participated, whereof 144 (3%) were seropositive at baseline. We analysed 107’820 questionnaires with a median follow-up of 7.9 months. Median number of answered questionnaires was similar (24 *vs*. 23 per person, P=0.83) between those with and without positive baseline serology. Among 2’713 HCW with ≥1 SARS-CoV-2 test during follow-up, 3/67 (4.5%) seropositive individuals reported a positive result (one of whom asymptomatic), compared to 547/2646 (20.7%) seronegative participants, 12 of whom asymptomatic (risk ratio [RR] 0.22; 95% confidence interval [CI] 0.07 to 0.66). Seropositive HCWs less frequently reported impaired olfaction/taste (6/144, 4.2% vs. 588/4674, 12.6%, RR 0.33, 95%-CI: 0.15-0.73), chills (19/144, 13.2% vs. 1040/4674, 22.3%, RR 0.59, 95%-CI: 0.39-0.90), and limb/muscle pain (28/144, 19.4% vs. 1335/4674, 28.6%, RR 0.68 95%-CI: 0.49-0.95). Impaired olfaction/taste and limb/muscle pain also discriminated best between positive and negative SARS-CoV-2 results.

**Conclusions:** Having SARS-CoV-2 anti-nucleocapsid antibodies provides almost 80% protection against SARS-CoV-2 re-infection for a period of at least eight months.

## INTRODUCTION

Effective and durable host immunity directed against severe acute respiratory syndrome (SARS-CoV-2) is key to the long-term control of the current Coronavirus Disease 2019 (COVID-19) pandemic. In consequence, the degree and duration of protection against re-infection in those with specific antibodies against SARS-CoV-2 are currently being debated [1]. Documented cases of reinfection (mean interval between infections was 106 days) are increasing and alternative avenues for immunity to SARS-CoV-2 have been proposed [2]. However, recent evidence suggests that neutralizing antibodies against SARS-CoV-2 are stably detectable for at least 9 months and offer protection against clinically relevant reinfection [3–5]. The most compelling evidence comes from a UK study, where - among 12’000 healthcare workers (HCWs) with a follow-up of 6 months - those with detectable anti-spike antibodies at baseline were less likely to have SARS-CoV-2 detected in a subsequent nasopharyngeal swab [4]. However, this study has not particularly assessed the frequency of COVID-19 specific symptoms among participants.

This HCW cohort study prospectively evaluated the risk of SARS-CoV-2 infection and the occurrence of COVID-19 symptoms among participants with and without SARS-CoV-2 anti-nucleocapsid antibodies at baseline.

## METHODS

### Study design

We initiated a prospective cohort study in 23 healthcare institutions in Northern and Eastern Switzerland. HCW with and without patient contact comprised the study population. Study approval was granted by the ethics committee of Eastern Switzerland (#2020-00502). Baseline results have been previously reported [6].

Upon study inclusion, participants provided blood for baseline serology. Subsequently, participants were tested through nasopharyngeal swabs (NPS) as soon as they experienced any COVID-19 compatible symptoms such as fever and/or the presence of any respiratory symptom (i.e. shortness of breath, cough, or sore throat). This symptom-based screening strategy was routinely implemented outside the study protocol in all participating institutions according to the recommendations of the Federal Office of Public Health. Also, HCWs residing in a bordering region of Austria or Germany were repetitively tested, irrespective of symptoms.

Via web-based questionnaire, participants responded to questions on demographics and occupation at baseline [6]. Participants were then prospectively followed and reminded by email and/or SMS to complete weekly web-based questionnaires. These requested data on COVID-19 compatible symptoms (syndromic surveillance) and date/result of any SARS-CoV-2 test (PCR/rapid antigen test). Participants were included up to the week where they reported having received their first dose of any SARS-CoV-2 vaccine or up to the end of the observation period, whichever came first.

### SARS-CoV-2 diagnostics

Details of serology testing at baseline are described elsewhere[6]. In brief, venous blood samples were analysed with an electro-chemiluminescence immunoassay (ECLIA, Roche Diagnostics, Rotkreuz, Switzerland, detection of total antibodies directed against the nucleocapsid-(N)-protein of SARS-CoV-2) [7]. Participants were informed about their individual test result.

Detection of SARS-CoV-2 from NPS was made by polymerase chain reaction (PCR) or rapid antigen test, depending on the method used in the participating institutions. To verify the completeness and accuracy of self-reported nasopharyngeal results, all self-reported positive tests and a random sample of negative test results were cross-checked with the database of the division of occupational health for a subgroup of HCWs from the largest participating institution.

### Data analysis

For the primary analysis, we compared the proportion of HCWs reporting at least one positive test result between the initially seropositive and seronegative individuals. This analysis was performed i) with tested HCWs only and ii) with all (tested and non-tested) HCWs as denominator. Risk ratios (RR) and corresponding 95% confidence interval (CI) were calculated. Furthermore, we compared the proportion of HCWs reporting at least one nasopharyngeal SARS-CoV-2 test during follow-up and, among these, the mean number of tests reported per person. Two sample proportion tests or Wilcoxon rank sum tests were used for this analysis.

Because previously seropositive participants might be tested less frequently than seronegative HCW, we also compared the frequency of self-reported symptoms according to serostatus at baseline among all study participants. For this analysis, questionnaires submitted within 2 weeks from baseline serology were excluded to avoid the detection of symptoms associated with episodes, which had started before baseline. For each participant, we summarized a symptom as present if reported in any of the submitted questionnaires, and absent otherwise; risk ratios (and 95% CI) and proportion tests were calculated for each symptom.

To assess specificity of symptoms regarding SARS-CoV-2 infection, we compared the frequency of symptoms between episodes with positive and negative SARS-CoV-2 test results. Only HCWs symptomatic at test time were included. Symptoms reported together with the test result and those reported in the previous and following questionnaires were linked to the respective episode. If a participant reported several swabs with the same result, only the first positive and/or the first negative swab were considered for this analysis. Thus, a participant contributed a maximum of one negative and one positive episode. In participants reporting both positive and negative tests, only negative tests preceding the first positive test by at least two weeks were included as a negative episode; any negative tests following a positive test were ignored. Odds ratios (OR) and 95% CIs were estimated for symptom specificity along with Fisher’s exact tests. Analyses were performed with R statistical software, version 4.0.2.

## RESULTS

We recruited HCW (n=4’818) between 22 June and 20 October 2020 (**Table 1**). At baseline, 144 (3%) participants were seropositive. Participants were followed until 9 March 2021, equalling a median follow-up of 7.9 months (interquartile range [IQR] 6.7-8.1 months). We received 107’820 weekly questionnaires from these 4’818 participants, corresponding to a response rate of 0.7 diaries per person and week. The median number of symptom diaries submitted before vaccination was 24 questionnaires (IQR 14-29) for initially seropositive and 23 (IQR 15-29) for seronegative participants (P=0.83).

**Table 1.** Distribution of baseline characteristics among the study participants, and distribution of SARS-CoV-2 serostatus for each level of the factors.

|  | Total N | Seropositive<br>(N and %) | Seronegative<br>(N and %) |
| --- | --- | --- | --- |
|  | N=4818 | N=144 | N=4674 |
| Gender |  |  |  |
| Female | 3763 | 108 (2.9%) | 3655 (97.1%) |
| Male | 1010 | 36 (3.6%) | 974 (96.5%) |
| Undetermined / missing | 45 |  |  |
| Age in years, median (IQR) | 38.9 (30.1–49.8) | 35.8 (27.4–45.1) | 39.0 (30.3–49.8) |
| BMI (kg m<sup>-2</sup>), median (IQR) | 23.4 (21.3–26.1) | 24.0 (22.0–26.7) | 23.4 (21.2–26.1) |
| Comorbidity |  |  |  |
| No (none mentioned) | 3081 | 86 (2.6%) | 2995 (97.2%) |
| Yes | 1719 | 58 (3.4%) | 1661 (96.6%) |
| Missing | 18 |  |  |
| Profession |  |  |  |
| Nurse or medical assistant | 2282 | 88 (3.9%) | 2194 (96.1%) |
| Physician | 809 | 13 (1.6%) | 796 (98.4%) |
| Other | 1495 | 31 (2.1%) | 1464 (97.9%) |
| None / missing | 232 |  |  |
| Speciality |  |  |  |
| Internal Medicine | 1025 | 34 (3.3%) | 991 (96.7%) |
| Surgery/Orthopedics | 466 | 15 (3.2%) | 451 (96.8%) |
| Intensive care | 326 | 8 (2.5%) | 318 (97.5%) |
| Emergency department | 267 | 9 (3.4%) | 258 (96.6%) |
| Other | 611 | 15 (2.5%) | 596 (97.5%) |
| None / missing | 2124 |  |  |
| Patient contact (medical or administration) |  |  |  |
| No | 772 | 12 (1.6%) | 760 (98.4%) |
| Yes | 3759 | 119 (3.2%) | 3640 (96.8%) |
| Missing | 287 |  |  |
| Involved in AGP |  |  |  |
| No | 3312 | 90 (2.7%) | 3222 (97.3%) |
| Yes | 1488 | 54 (3.6%) | 1434 (96.4%) |
| Missing | 18 |  |  |

Abbreviations: IQR, Interquartile Range; BMI, Body Mass Index; AGP, Aerosol-Generating Procedure

A total of 2’713 individuals reported having at least one test (PCR/rapid antigen testing) performed during follow up. Seropositive participants were less likely (67/144, 47%) to undergo testing than seronegative participants (2’646/4’674, 57%) (P=0.02). Conversely, the mean number of tests per person (among those with at least one test) did not differ significantly between seropositive and seronegative HCWs (1.8 vs. 2.0 tests, P=0.33).

In total, 550 of 2’713 tested participants reported at least one positive test result during follow-up. Of 67 seropositive participants who underwent testing, only three (4.5%) received at least one positive test result during follow-up, compared with 547 (20.7%) of the 2’646 seronegative tested participants. (**Figure 1**). This translates into a RR of 0.22 (95%-CI: 0.07 to 0.66, P=0.002) for a positive SARS-CoV-2 test after positive baseline serology.

**Figure 1.**
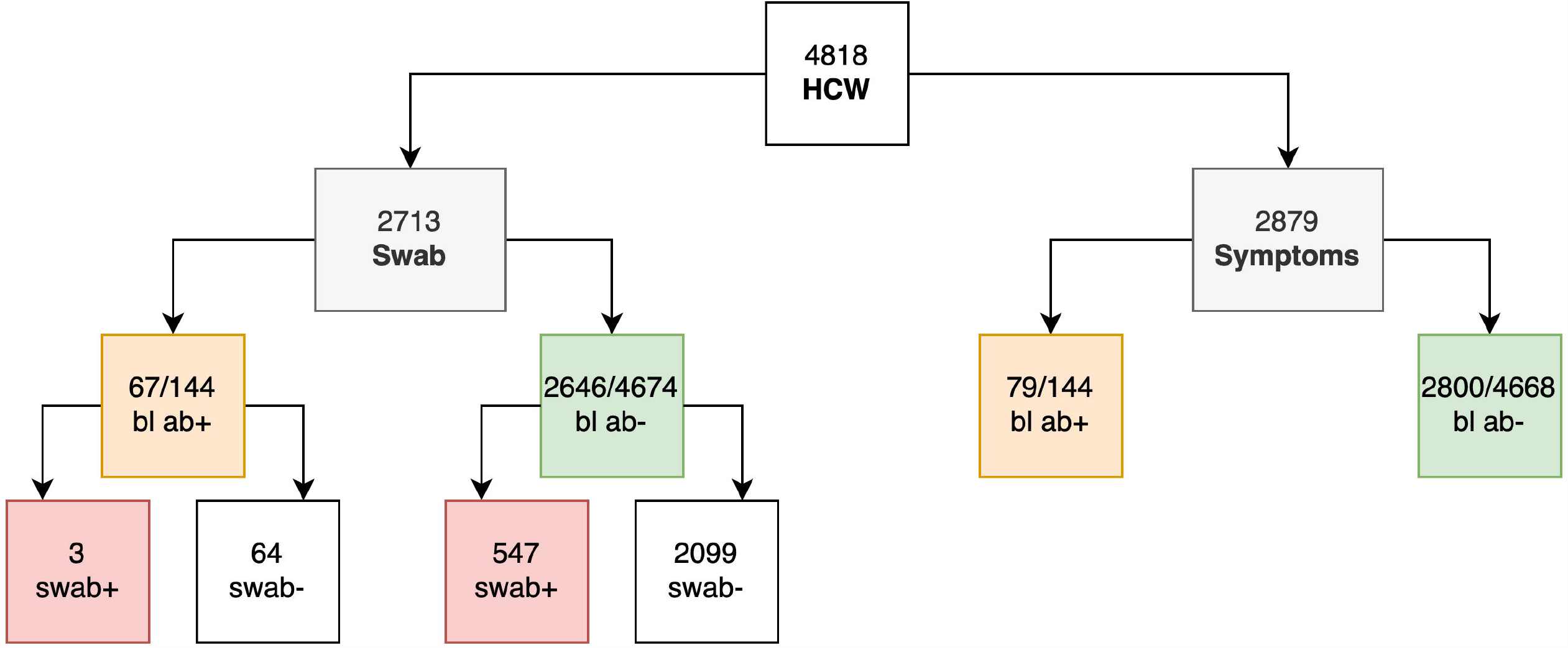
Flow chart, illustrating the distribution of health workers who had at least one nasopharyngeal swab for SARS-CoV-2, and of those who reported any COVID-19 compatible symptoms, according to diagnostic test results. **Abbreviations**: **ab** = antibody, **bl** = baseline, **HCW** = health care worker, **+** = positive, **-** = negative **Colours**: grey = outcomes, yellow = baseline seropositive, green = baseline seronegative, red = nasopharyngeal swab positive in minimally one swab

Considering all (tested and non-tested) HCWs, the corresponding RR was 0.18 (95%-CI: 0.06 to 0.55, P<0.001). The three cases with presumable re-infection after positive baseline serology were all diagnosed in January 2021 after a follow-up (i.e. time from baseline serology to second positive SARS-CoV-2 test) of 198, 200, and 220 days. One of the three HCWs was asymptomatic at time of re-infection. For detailed characteristics, see **Table 2**.

**Table 2.** Characteristics of three participants with positive SARS-CoV-2 nasopharyngeal swab after positive baseline serology.

|  | HCW 1 | HCW 2 | HCW 3 |
| --- | --- | --- | --- |
| Sex | Female | Female | Male |
| Age range at re-infection | 50-55 | 50-55 | 40-45 |
| Blood group | Unknown | B | B |
| Comorbidities | None | Pollen allergy | Pollen allergy |
| Profession | Nurse (assistant) | Nurse (assistant) | Nurse (assistant) |
| Patient contact | Administrative | Administrative / caring | Administrative |
| First positive PCR | March 2020 | Date unknown | March 2020 |
| Date baseline serology | July 2020 | July 2020 | June 2020 |
| Second positive test (method) | January 2021 (PCR) | January 2021 (PCR) | January 2021 (PCR) |
| Days baseline to second episode | 198 | 200 | 220 |
| Days first to second episode | 297 | Unknown | 314 |
| Negative tests between first and second episode | None | None | One |
| Symptoms second test | Sore throat, coryza, headache, limb/muscle pain, eye irritation, weakness | None | Headache, dizziness, diarrhea, shivering, limb/muscle pain, eye irritation, weakness |
| Exposure | Patients | Patients | Patients, household |
| Number or household members | One | One | One |
| COVID-19 vaccine | None | None | First dose in week of positive test |

**Abbreviations:** HCW, Healthcare Worker; PCR, Polymerase Chain Reaction; COVID-19, Coronavirus Disease-2019

Among 4’812 participants (with 101’233 questionnaires submitted at least 2 weeks after baseline), 2’879 HCW reported at least one symptom during follow-up. Symptoms were reported by 79/144 (54.9%) of the initially seropositive participants and by 2’800/4’668 (60.0%) of the initially seronegative participants (P=0.25) (**Figure 1**). The total number of reported symptoms was not significantly different in baseline seropositive compared to seronegative individuals (median of 5 *vs*. 6 symptoms, P=0.30). Out of 15 different symptoms, ten were reported less frequently by participants who were seropositive at baseline, although the difference was statistically significant only for impaired olfaction/taste (RR 0.33, 95%-CI: 0.15-0.73, P=0.004), chills (RR 0.59, 95%-CI: 0.39-0.90, P=0.01), and limb/muscle pain (RR 0.68 95%-CI: 0.49-0.95, P=0.02) (**Figure 2, Table 3**).

**Figure 2.**
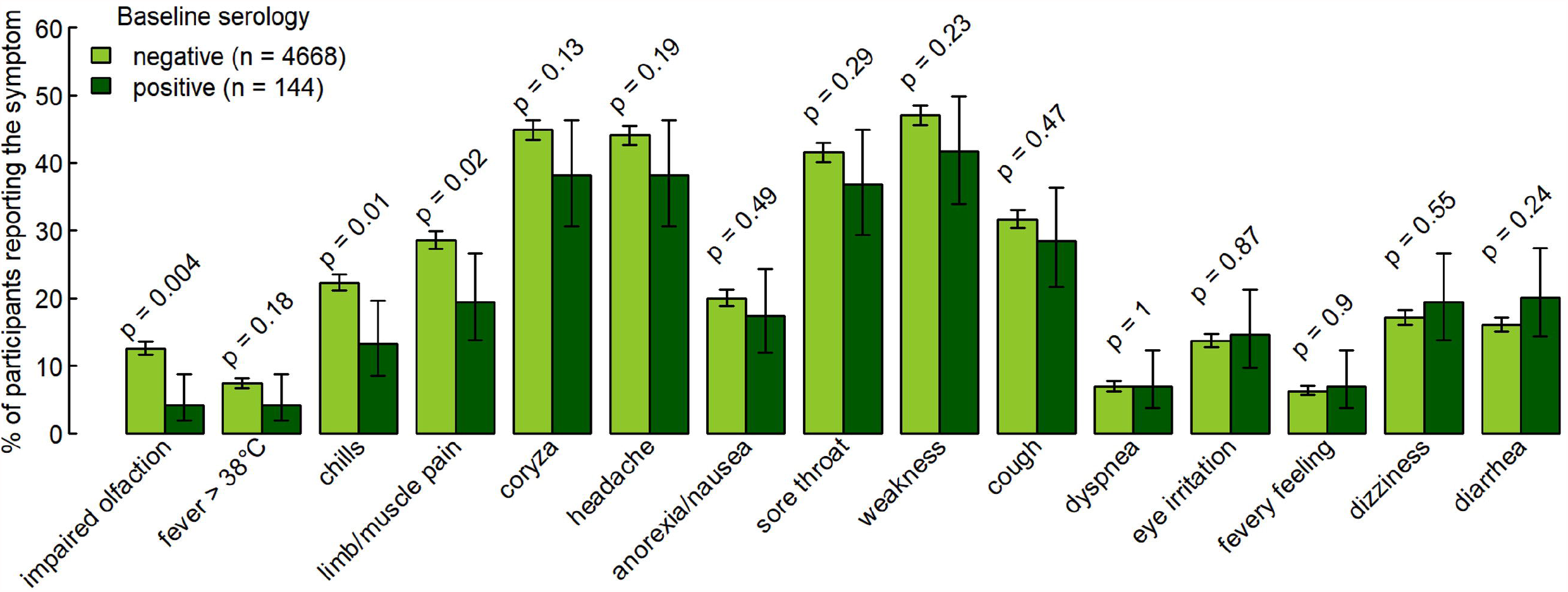
Frequency of symptoms during a median follow-up of 7.9 months based on 101’233 weekly diaries from participants with (n=144) and without (n=4’668) specific SARS-CoV-2 antibodies at baseline. Symptoms are sorted by increasing risk ratio. Error bars show 95% Wilson confidence intervals. P-values calculated by two-sample proportion test.

**Table 3.** Frequency of individual symptoms (n and % of participants who reported it at least once during study participation more than 2 weeks after baseline and prior to vaccination) in relation to baseline serology. Symptoms are sorted by increasing risk ratio (RR).

| Symptom | N (%) of HCWs reporting a symptom |  | RR (95% CI) | *p*-value<br>prop.test* | *p*-value<br>adjusted** |
| --- | --- | --- | --- | --- | --- |
|  | Seronegatives | Seropositives |  |  |  |
|  | N=4668 | N=144 |  |  |  |
| Olfaction/taste impaired | 588 (12.6%) | 6 (4.2%) | 0.33 (0.15-0.73) | 0.004 | 0.001 |
| Fever > 38°C | 348 (7.5%) | 6 (4.2%) | 0.56 (0.25-1.23) | 0.185 | 0.101 |
| Chills | 1040 (22.3%) | 19 (13.2%) | 0.59 (0.39-0.90) | 0.013 | 0.005 |
| Limb/muscle pain | 1335 (28.6%) | 28 (19.4%) | 0.68 (0.49-0.95) | 0.021 | 0.01 |
| Coryza/nasal congestion | 2094 (44.9%) | 55 (38.2%) | 0.85 (0.69-1.05) | 0.134 | 0.083 |
| Headache | 2058 (44.1%) | 55 (38.2%) | 0.87 (0.70-1.07) | 0.187 | 0.126 |
| Anorexia/Nausea | 935 (20%) | 25 (17.4%) | 0.87 (0.60-1.24) | 0.494 | 0.393 |
| Sore throat | 1940 (41.6%) | 53 (36.8%) | 0.89 (0.71-1.10) | 0.292 | 0.207 |
| Weakness | 2196 (47%) | 60 (41.7%) | 0.89 (0.73-1.08) | 0.235 | 0.162 |
| Cough | 1478 (31.7%) | 41 (28.5%) | 0.90 (0.69-1.17) | 0.471 | 0.37 |
| Dyspnea | 326 (7%) | 10 (6.9%) | 0.99 (0.54-1.82) | 1 | 0.962 |
| Eye irritation | 641 (13.7%) | 21 (14.6%) | 1.06 (0.71-1.59) | 0.866 | 0.805 |
| Fevery feeling | 295 (6.3%) | 10 (6.9%) | 1.10 (0.60-2.02) | 0.897 | 0.789 |
| Dizziness | 801 (17.2%) | 28 (19.4%) | 1.13 (0.81-1.59) | 0.546 | 0.509 |
| Diarrhea | 751 (16.1%) | 29 (20.1%) | 1.25 (0.90-1.74) | 0.236 | 0.222 |

**Abbreviations:** RR, Risk Ratio; CI, Confidence Interval
\* two-sample proportion test
\*\* based on a logistic regression model including the log-transformed number of questionnaires submitted as covariate

For the analysis of symptom specificity, we included 532 episodes with positive and 1’527 with negative test results. Almost all (14 out of 15) symptoms were less frequently reported during episodes with a positive NPS (compared to episodes with negative test). The symptoms which discriminated best between episodes with positive and negative SARS-CoV-2 nasopharyngeal test were impaired olfaction/taste (OR 22.2 95%-CI: 17.1-29.1, P<0.001), limb/muscle pain (OR 4.7 95%-CI: 3.8-5.9, P<0.001), and weakness (OR 4.4 95%-CI: 3.1-6.4, P<0.001) (**Figure 3, Table S1**).

**Figure 3.**
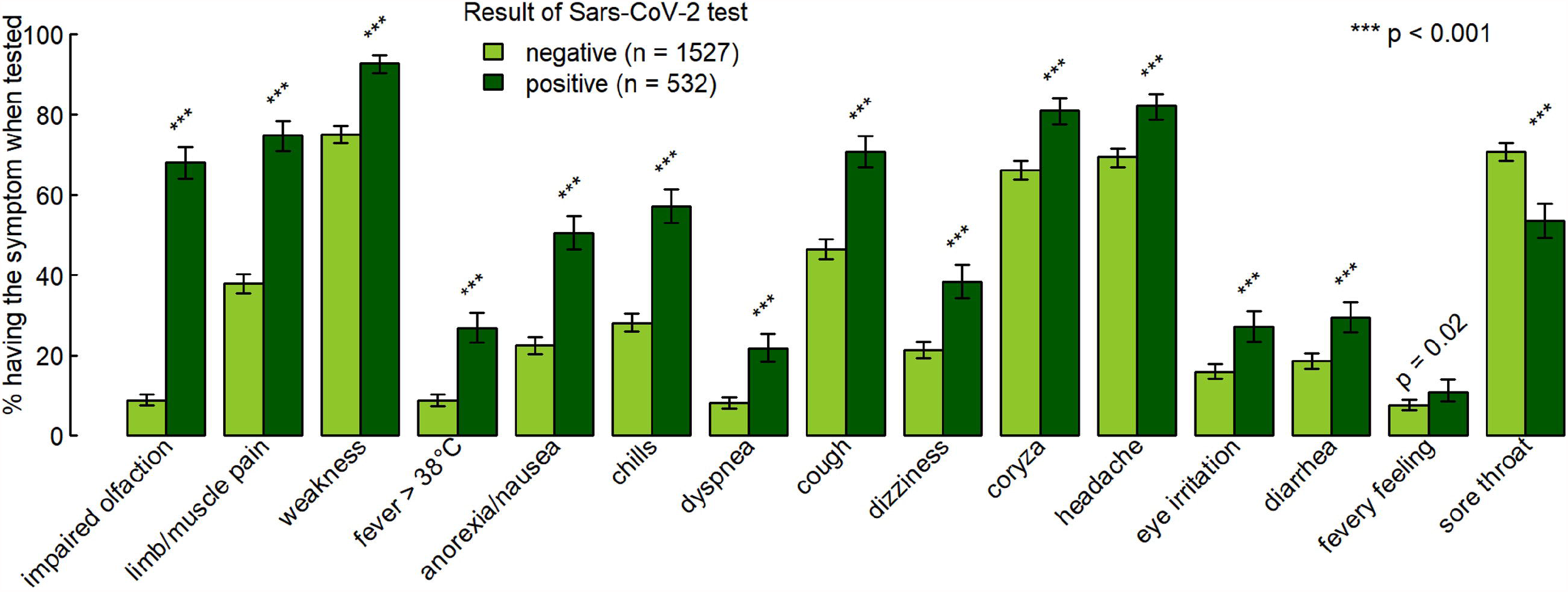
Percentage of participants reporting individual symptoms at the time of either negative or positive nasopharyngeal swabs, with 95% Wilson confidence intervals and with *p*-values from Fisher’s exact tests. Symptoms are sorted by decreasing odds ratio (OR) for occurrence together with a positive swab (see also Table S1).

For validation of swab results, we cross-checked self-reported test results for a subgroup of participants (from the largest participating institution). We found that 150 out of 174 presumable positive tests were indeed documented in the database of the division of occupational medicine. The remaining HCWs most likely having had a positive test outside of their working place. On the other hand, none of the randomly selected 175 HCWs reporting only negative test results was found to have a positive result in the database.

## DISCUSSION

In this prospective cohort of almost 5’000 HCW followed during the second wave in Switzerland, we demonstrate that the presence of anti-nucleocapsid SARS-CoV-2 antibodies at baseline not only reduced the risk of positive nasopharyngeal SARS-CoV-2 tests, but also the occurrence of COVID-19 specific symptoms such as loss of smell and limb or muscle pain. The follow-up of almost eight months, the large sample size, the systematic collection of symptoms, and the excellent questionnaire response rate are among the strengths of the study.

These results add to the mounting evidence that specific antibodies protect against subsequent SARS-CoV-2 infection. In the study of Lumley *et al*., the adjusted incidence rate ratio for seropositive HCW to have a positive PCR (median follow-up 6 months) was 0.11 compared to those without antibodies [4]. Within a cohort of over 3.2 million US patients, Harvey *et al*. found a ratio of 0.10 of positive PCRs among those with *vs*. those without positive antibody test at baseline (for tests performed >90 days after baseline) [8]. Among over 43’000 people with a positive antibody test (median follow up 4 months) from Qatar, the estimated efficacy of natural infection against reinfection was above 90% [9]. Also, data from a population-based study (>500’000 people) conducted in Denmark suggest that protection after natural SARS-CoV-2 infection was 80% after 6 months (but only 47% for adults aged 65 years or older) [10]. A retrospective propensity-score matched cohort study from Western Switzerland found a 94% reduction in the hazard of having a positive SARS-CoV-2 test for seropositive individuals [11]. In a large HCW cohort from England, a previous history of SARS-CoV-2 infection was associated with a 84% lower risk of infection (median follow-up 7 months) [12]. Likewise, in a university student population from the US undergoing repeat mandatory testing, a 84% protection from SARS-CoV-2 infection was found [13]. Although our median follow-up of eight months is among the longest compared with other studies, our point estimate (RR 0.22) for protection from reinfection with SARS-CoV-2 is perfectly in line with these results. Our findings are further supported by a recent modelling study of the progression over 250 days of neutralising antibodies following infection or vaccination and their protective role against symptomatic SARS-CoV-2 infection [14].

Previous studies have mainly looked at the incidence of confirmed SARS-CoV-2 infection to assess the risk of re-infection. This approach has the inherent limitation that people who are seropositive might be less likely to undergo testing, as seen in our data. In addition to testing for SARS-CoV-2, we therefore used a second approach to assess the protective effect of seropositivity on subsequent COVID-19. Using weekly symptom frequency as proxy for COVID-19 allows comparing incidence of COVID-19 irrespective of whether patients underwent testing or not. Of course, this signal is being diluted by infections caused by other respiratory viruses, especially for symptoms like coryza, sore throat, cough or fever. However, certain symptoms such as loss of taste or smell and myalgias have already been shown by others to be more specific for COVID-19 [15,16]. The fact that exactly these symptoms occurred less frequently among those with antibodies at baseline supports our finding of a reduced risk for SARS-CoV-2 re-infection in this group. A limitation of this approach is that some participants might suffer from persisting COVID-19 symptoms (i.e. long-COVID). In particular loss of smell has been shown to persist for several weeks in a certain proportion of COVID-19 patients [17]. If we had excluded seropositive participants who already reported this symptom at baseline, the effect would have even been more pronounced.

Worryingly, re-infections with phylogenetically different SARS-CoV-2 strains are increasingly being reported [18]. A recent report documented severe reinfection with the “new variant” VOC-202021/01 eight months after documented primary infection, even in the presence of anti-nucleocapsid antibodies and in the absence of overt immunosuppression [19]. We also observed three nurse assistants with presumable SARS-CoV-2 re-infection approximately 300 days after first infection and about 6 months after documented seroconversion. Of note, one of the nurses did not report any symptoms at time of the test. Although we do not have any sequencing data for these particular viral strains, reinfections occurred in January 2021, when the proportion of the B.1.1.7 variant was estimated to account for less than 20% of all SARS-CoV-2 isolates in Switzerland [20]. Continuing follow-up of our and other cohorts will reveal the long-term protective effect of specific antibodies against SARS-CoV-2 and its emerging variants. Another open research question regards the long-term protective effect of vaccine-induced immunity, which might be higher compared to natural infection [14,21].

A limitation of our study is the fact that nasopharyngeal SARS-CoV-2 testing was not routinely performed. Therefore, asymptomatic carriage of SARS-CoV-2 cannot be excluded in those with antibodies at baseline. We also determined SARS-CoV-2 anti-nucleocapsid antibody titers and not anti-spike antibodies, which have been shown to correlate better with virus neutralization. Post SARS-CoV-2 infection, anti-N antibodies are detected equally[22] or even more frequently[23] than anti-S antibodies. We therefore suggest that our results represent, or somewhat underestimate the true protective effect mediated by anti-S antibodies. Swab results and symptoms were self-reported. Because validation of some of the swab results showed mostly consistent results, we consider these self-reported data to be highly reliable. Furthermore, we cannot definitely confirm that the three seropositive HCWs with positive SARS-CoV-2 NPS were indeed re-infected with a new strain. However, the long latency between the episodes, new onset of symptoms (two cases), and a negative PCR between episodes (one case) strongly support our hypothesis of re-infection (rather than persistence of viral RNA for more than 6 months). Another shortcoming of this study is that SARS-CoV-2 specific cellular immunity was not evaluated. This is mainly due to the fact that these measurements are still very time-consuming and cost-intensive in a large population. Thus, data are scarce so far. However, specific T-cells most likely also contribute decisively to protection against SARS-CoV-2 [24]. Immunity mediated by specific T cells can be present even if there have never been signs of disease and antibodies are absent [25,26]. In consequence, measuring antibodies alone, such as in our study, underestimates protection against COVID-19 in a population.

We conclude that anti-nucleocapsid antibodies convey an approximately 80% protection against symptomatic SARS-CoV-2 infection, at least for a period of 8 months and in a setting where “new variant” mutations were not widely present at the end of follow-up. Syndromic surveillance for specific COVID-19 symptoms allows estimating the probability of SARS-CoV-2 re-infection irrespective of whether participants undergo testing or not.

## Data Availability

Data are available from the corresponding author upon reasonable request.

## Acknowledgments

We would like to warmly thank the large number of employees of the participating health care institutions who either took part in this study themselves or supported it. Furthermore, we thank the laboratory staff for shipment, handling and analysis of the blood samples. In particular, we acknowledge the organizational core team Simone Kessler and Susanne Nigg, who kept all strings between the participating centres and the laboratory and without whom this study would not have been possible.

## Author’s contributions

All authors contributed to the conceptualization of the study. PK and CRK supervised the study. PK, OL, TE, PV and CRK were responsible for data curation. TE was responsible for project administration. PK, MS, DF, AB, EL, CM, PR, RS, DV, BW, UB, LR and AF contributed to the investigations. LR and CRK provided laboratory resources. SG was responsible for the formal analysis and data visualizations. PK, PV and CRK were responsible for funding acquisition. PK and CRK wrote the original draft, which was critically reviewed and edited by all authors.

