## Supplemental Table 1 for "Impact of baseline SARS-CoV-2 antibody status on syndromic surveillance and the risk of subsequent Covid-19 – a prospective multicentre cohort study"

**SUPPLEMENTS**

Table S1. Frequency of individual symptoms around the time of a nasopharyngeal swab in relation to the test result (negative or positive), considering only participants who reported at least one symptom at this time. Symptoms are sorted by decreasing odds ratio (OR).

| Symptom | n (%) of HCWs reporting a symptom | | OR (95% CI)* | *p*-value |
|---|---|---|---|---|
| | negative swab N=1527 | positive swab N=532 | | Fisher |
| Olfaction/taste impaired | 134 (8.8%) | 362 (68%) | 22.07 (17.03-28.79) | <0.001 |
| Limb/muscle pain | 578 (37.9%) | 398 (74.8%) | 4.87 (3.89-6.13) | <0.001 |
| Weakness | 1146 (75%) | 494 (92.9%) | 4.32 (3.03-6.3) | <0.001 |
| Fever > 38°C | 133 (8.7%) | 142 (26.7%) | 3.81 (2.91-5) | <0.001 |
| Anorexia/nausea | 342 (22.4%) | 269 (50.6%) | 3.54 (2.86-4.39) | <0.001 |
| Chills | 429 (28.1%) | 304 (57.1%) | 3.41 (2.77-4.21) | <0.001 |
| Dyspnea | 123 (8.1%) | 115 (21.6%) | 3.15 (2.36-4.19) | <0.001 |
| Cough | 708 (46.4%) | 377 (70.9%) | 2.81 (2.26-3.5) | <0.001 |
| Dizziness | 326 (21.3%) | 204 (38.3%) | 2.29 (1.84-2.85) | <0.001 |
| Coryza/nasal congestion | 1011 (66.2%) | 431 (81%) | 2.18 (1.7-2.8) | <0.001 |
| Headache | 1059 (69.4%) | 437 (82.1%) | 2.03 (1.58-2.63) | <0.001 |
| Eye irritation | 242 (15.8%) | 144 (27.1%) | 1.97 (1.54-2.51) | <0.001 |
| Diarrhea | 283 (18.5%) | 156 (29.3%) | 1.82 (1.44-2.3) | <0.001 |
| Fevery feeling | 115 (7.5%) | 58 (10.9%) | 1.5 (1.06-2.12) | 0.02 |
| Sore throat | 1080 (70.7%) | 285 (53.6%) | 0.48 (0.39-0.59) | <0.001 |

* estimated through the procedure implemented in Fisher's exact test
